# Direct and indirect effects of COVID-19 on infant bronchiolitis: a population-wide registry study from Northern Italy

**DOI:** 10.64898/2026.09.22.26363663

**Authors:** Valeria Nazzari, Albert Navarro Gallinad, Laura Ghirardi, Claudia Gianbartolomei, Anna Maria Paganoni, Azucena Bardají, Luisa Zuccolo

**Affiliations:** Health Data Science Centre, Human Technopole, Milan, Italy; MOX, Department of Mathematics, Politecnico di Milano, Piazza Leonardo da Vinci 32, Milan, 20133 Italy; ISGlobal, Barcelona, Spain; Centro de Investigação em Saúde de Manhiça (CISM), Manhiça, Mozambique; Consorcio de Investigación Biomédica en Red de Epidemiología y Salud Pública (CIBERESP), Madrid, Spain; Facultat de Medicina i Ciències de la Salut, Universitat de Barcelona (UB), Barcelona, Spain; MRC Integrative Epidemiology Unit at the University of Bristol, Bristol, UK

## Abstract

**Background:** COVID-19 pandemic modified trends of infant bronchiolitis, leading cause of emergency room (ER) attendance in infants in high income countries, primarily due to Respiratory Syncytial Virus. Understanding recent pandemic-related epidemiological shifts could inform affordable prevention efforts to improve sustainability and coverage of prophylactic treatments beyond maternal vaccines and infant monoclonal antibodies.

**Methods:** We estimated incidence rates (IR) of bronchiolitis ER presentations in 0-6month olds in 2012-2023 in a region-wide cohort of almost 1 million births using administrative health data from Lombardy, Italy. To assess potential contributors, we investigated direct and indirect COVID-19-related factors, including disease severity, shape of the epidemic peak, parental health seeking behaviour and vaccine coverage. We estimated COVID-19 maternal vaccination associations using mothers-infants data linkages and multivariable-adjusted logistic regression.

**Findings:** Compared to pre-pandemic levels, IRs doubled in 2021-2023 (from 2 to ≥ 4/100 person-time). The surge could not be explained by changes in infant age at ER visit, clinical severity or in the shape of the epidemic peak. Adjusted models showed little support for a role of infant routine or maternal COVID-19 vaccination on bronchiolitis episodes (respectively hazard ratio 1.09, 95%CI: 0.98–1.22 and odds ratio 0.91, 95%CI: 0.76–1.09).

**Interpretation:** The post-pandemic increase in infant bronchiolitis is likely driven by multiple epidemiological changes rather than a single factor. As maternal RSV vaccination and infant RSV monoclonal antibodies are introduced and scale up, identifying these mechanisms will be critical to maximise their public health impact, inform complementary prevention strategies, and improve preparedness for future RSV epidemics.

## Introduction

Respiratory Syncytial Virus (RSV) is a major cause of lower respiratory tract infections in young children globally, and the major cause of emergency room (ER) visits and hospital admissions in infants in high-income countries^1^. The typical presentation is bronchiolitis, an acute lower respiratory tract infection of the small airways with approximately 85% of infant cases attributed to RSV^2^. The COVID-19 pandemic dramatically altered the seasonal pattern of infant bronchiolitis worldwide^3^. Before the pandemic, hospitalizations showed distinct seasonal trends peaking in winter^4^; while during 2020-2021, RSV-related hospitalizations declined, likely due to COVID-19 related lockdown and public health measures^5^. In 2021-2022, RSV cases spiked globally, with infant bronchiolitis admissions doubling compared with pre-pandemic levels across several countries^6^. In Italy, reports on recent bronchiolitis trends have been inconsistent and often limited by local coverage or reliance on absolute case numbers rather than population-based rates^7–10^. Consequently, the population-level magnitude of these changes and, importantly, the factors underlying them remain unclear.

Current prevention options for RSV and infant bronchiolitis include recently approved and highly effective prophylactic treatments aimed at both infants (the monoclonal antibody Nirsevimab^11^) and pregnant women (the RSV bivalent prefusion F protein-based vaccine^12^). These are not universally available, including in high-income settings such as Europe^13^, due to their high costs. It is therefore highly desirable, if not necessary, to identify additional, more affordable public health measures which could reverse the post-pandemic sharp and sudden increase in infant bronchiolitis cases and anticipate future RSV outbreaks.

Several explanations have been put forward for the recent rise in infant bronchiolitis, although their relevance and relative contribution remain unclear^14^. These can be categorised as direct and indirect impact of COVID-19. Among the former, are long-term alterations in immune regulation following SARS-CoV-2 infection resulting in heightened susceptibility to viral infections and therefore RSV^15^. Among the latter, suggestions include pandemic-related reduced primary care visits (with consequent increased presentation of non-severe cases at emergency care settings), increased healthcare seeking behaviour on behalf of parents of young children (with consequent multiple hospital visits for the same viral episode), delayed vaccination appointments (with consequent older age at full immunisation protection), and generally lower uptake of maternal and infant vaccines (with consequent increased numbers of infants at risk of infections). During lockdowns, disrupted postnatal care could have led to lower breastfeeding rates^16^ and reduced population immunity following lower RSV circulation may have increased susceptibility among infants^17,18^. In this context, in-depth epidemiological analyses of time trends and pandemic-associated risk factors could provide novel insights into the changing epidemiology of RSV in infants, the landscape of emerging preventative interventions and their optimal deployment.

Here, we model changes in incidence rates of ER admissions for bronchiolitis in Italian infants between 2012-2023. We explore the potential explanations behind recent significant changes, including direct and indirect effects of COVID-19. After ruling out artefacts such as shifts in population demographics or in the shape of epidemic peak, we conduct a range of analyses to identify key drivers of these trends. We investigate the direct role of the new SARS-CoV2 pathogen by comparing bronchiolitis risk in infants born to mothers with and without COVID-19 vaccine protection during pregnancy. We then investigate indirect COVID-19 impacts, by examining changes in bronchiolitis rates due to the presumed consequences of changes in healthcare seeking behaviour or healthcare provision including non-RSV-specific vaccinations (infant: hexavalent diphtheria, tetanus, acellular pertussis, inactivated poliovirus, hepatitis B, and Haemophilus influenzae type b (DTaP-IPV-HepB-Hib) and pneumococcus vaccines; mother: DTaP vaccine).

## Materials and Methods

### Data source

We use linked administrative data on healthcare utilisation covering the entire population of in Lombardy, a Northern-Italian region (population approximately 10 million people^19^). The database includes demographic information, hospital discharges, ER visits, vaccination, drugs dispensations, and certificates of delivery assistance (CeDAP), the equivalent of a medical birth registry. Datasets are linkable through a unique pseudo-anonymized individual identifier, including mother-child linkage using pregnancy-specific identifiers. The dataset available includes women registered in the regional registry and residing as of December 1, 2019, born between 1973 and 2008, and children born between January 1, 2012, and June 30, 2023. Additional details on data sources and study variables are provided in Table S1.

### Paediatric and Maternal-Paediatric cohort

We define three distinct cohorts: (i) a dynamic paediatric cohort of 0-6 months old infants residing in Lombardy between May 2012-May 2023, for descriptive trend analyses; (ii) a restricted paediatric cohort including only infants aged 0-6 months born in Lombardy between June 2021-November 2022, for hypothesis-testing analyses (direct effects of COVID-19); and (iii) a maternal-paediatric cohort of mother-infant pairs linked through CeDAP identifiers, for hypothesis-testing analyses (direct effects of COVID-19). In the maternal-paediatric cohort, inclusion criteria were pregnancies with last menstrual period on or after January 1^st^, 2021, maternal residence in Lombardy throughout the entire pregnancy and children residence in the region from birth until six months of age. Cohorts’ selection is shown in Figure 1.

**Figure 1:**
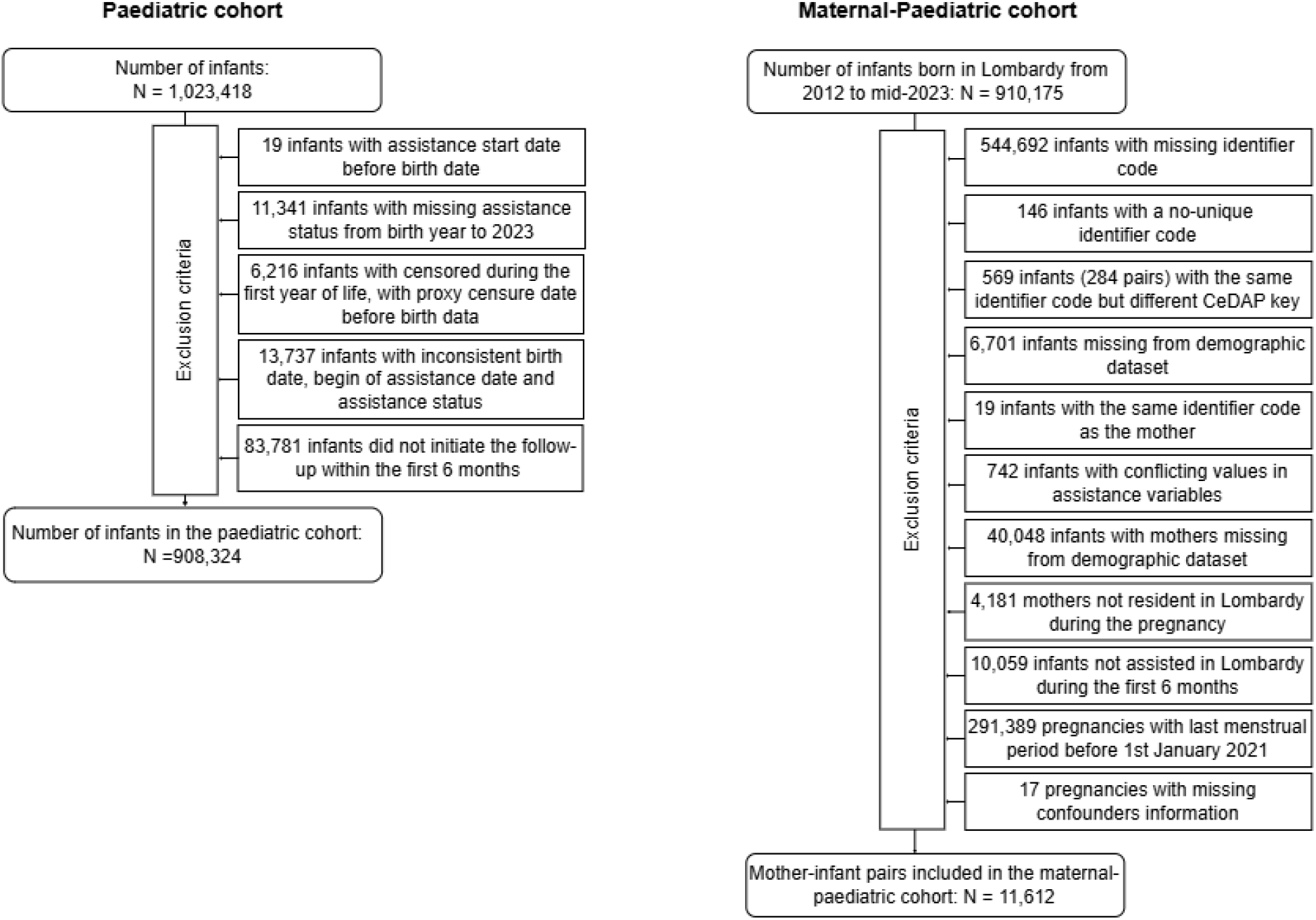
Flow diagram of exclusion ad inclusion criteria used to derive the paediatric and maternal-paediatric cohorts.

### Exposure, outcome and potential confounders

Bronchiolitis cases were identified from the Emergency Room dataset using ICD-9-CM code 466.1* (acute bronchiolitis), excluding records with missing primary diagnosis or inconsistent admission/discharge dates. DTaP-IPV-HepB-Hib, pneumococcal, COVID-19, and Tdap vaccinations were identified ATC codes J07CA09, J07AL02, J07BX03 (including the subsequent J07BN series^20^), and J07AJ52 respectively. Confounders included maternal demographic characteristics and clinical history derived from hospital discharge records, ER visits, and drug dispensations datasets (Table S2).

### Statistical analysis

#### Trend analysis

We estimated monthly bronchiolitis incidence rates among infants aged 0–6 months by dividing the number of emergency room visits by the corresponding person-time at risk within the paediatric cohort. These monthly rates were then used to construct a time series for trend analysis for visual inspection and quantification of epidemic peak height and shape.

#### Artefacts

To explore alternative explanations for potential trend changes (pre-post pandemic differences in epidemic peak height and shape), we first investigated potential artefacts. Time series were constructed from incidence rates, rather than total number of cases, allowing us to account for shifts in population demographics and to analyse changes in seasonal epidemic peaks. The latter was evaluated by calculating the overall seasonal incidence, operationalized as the Area Under the Time Series (AUTS) for each 12-month epidemiologic year (June to May).

#### COVID-19 direct effects

We used maternal COVID-19 vaccination during pregnancy as a proxy for prenatal SARS-CoV-2 immunity to investigate the potential role of SARS-CoV-2 in the development of bronchiolitis.

Within the maternal–paediatric cohort, we assessed whether this proxy measure was associated with the risk of bronchiolitis in offspring. Exposure was defined as receipt of at least one dose of a COVID-19 vaccine during pregnancy, while the outcome was defined as at least one ER visit for bronchiolitis during the infant’s first six months of life. We used logistic regression to estimate odds ratios (ORs) and 95% CIs. We fitted unadjusted, partially adjusted (categorical month-year of delivery and newborn sex.), and fully adjusted models. The fully adjusted model additionally included age at delivery, maternal birth country (Italy/abroad), domicile at delivery, employment and educational level, and indicators of maternal health status listed in Table S2. We conducted two sensitivity analyses. First, we restricted to first-born infants. Second, we investigated immune protection derived from a broader prenatal immunization, extending the analysis including maternal Tdap vaccination. Women who received neither COVID-19 nor Tdap vaccination served as the reference group, while exposed women were classified into three distinct categories: COVID-19 vaccination only, Tdap vaccination only, or both.

#### COVID-19 indirect effects

First, we explored whether changes in parental health seeking behaviour could explain the post-pandemic trend increases. This was done by examining the distribution of disease severity and accounting for multiple visits pertaining to the same disease episode. We evaluated severity by constructing a time series of ER visits that resulted in hospitalization, while visit frequency was assessed by applying a 15-day washout period between ER presentations to account for repeated visits during the same illness episode. We then investigated whether delayed immunisation schedules could play a role, by visually inspecting the yearly age distribution at ER presentation, calculated as the relative frequency of cases at each month of age for each 12-month period.

Finally, we evaluated the association with broader non-RSV specific immune protection in infants, to assess the likelihood that lower vaccine uptake in general could increase disease susceptibility. We examined whether early infants’ immunisation protects from bronchiolitis ER visits using a restricted paediatric cohort. Exposure was defined as the administration of at least one dose of the hexavalent or pneumococcal vaccines within the first 90 days of life, capturing the initiation of the primary vaccination schedule. To account for potential immortal time bias, we employed a Cox proportional hazard model with a time-dependent exposure to estimate the hazard ratio (HR) and 95% CIs, accounting for a 14-day induction period for development of an immune response. We fitted both an unadjusted model and an adjusted model that accounted for month and year of birth.

## Results

### Trend analysis

The paediatric cohort included 908,324 infants (0–6 months). Person-time decreased over the study period, reflecting a decline in the number of eligible infants in Lombardy (Figure S1). Between the 2012–2013 and 2019–2020 bronchiolitis seasons, incidence rate peaks of bronchiolitis remained stable at approximately 2 cases per 100 person-months, with a consistent seasonal peak occurring between December and January (Figure 2). During the 2020–2021 season, corresponding with COVID-19 lockdown measures, incidence rate dropped sharply, reaching nearly zero. In the subsequent 2021–2022 and 2022–2023 seasons, incidence rate of bronchiolitis approximately doubled compared to pre-pandemic levels, rising to around 4 cases per 100 person-months. The seasonal peak shifted earlier to November in 2021–2022, before returning to December 2022–2023.

**Figure 2:**
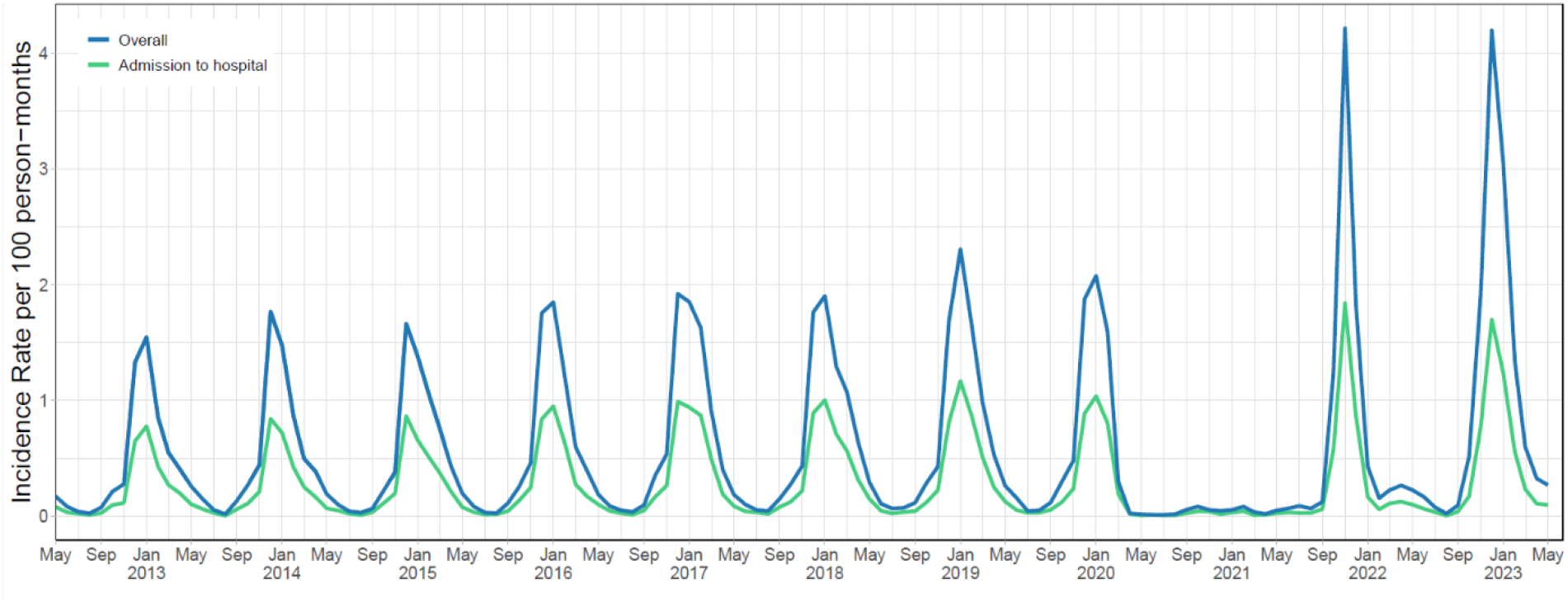
Monthly incidence rates of bronchiolitis emergency room visits and visits resulting in hospital admission among infants aged 0–6 months.

### Artefacts

Analysis of the AUTS estimates revealed an increase in cumulative bronchiolitis burden during the last two seasons, with a 50.5% higher mean seasonal burden compared with the pre-pandemic period (Table S3).

### COVID-19 direct effects

A total of 11,612 infants from 11,441 distinct mothers were included in the maternal–paediatric cohort. Among them, 6,744 infants (58.1%) were born to mothers who received at least one dose of a COVID-19 vaccine during pregnancy (exposed group). Vaccinated mothers were slightly older (mean age 34.2 vs. 33.3 years), more frequently born in Italy (78% vs. 68%), and more likely to be employed and highly educated (Table 1). During the follow-up period, 648 (5.6%) ER visits for bronchiolitis were recorded. Of these, 298 (46%) ER visits occurred among infants whose mothers received COVID-19 vaccination (exposed group), and 350 (54%) among those in the unexposed group. Based on the adjusted model, there was no evidence that COVID-19 vaccination during pregnancy decreased the risk of infant bronchiolitis ER visits (OR=0.91 [0.76– 1.09]). The inverse association observed in the unadjusted analysis was substantially attenuated after adjustment for month-year of delivery and infant sex, with little further change after full adjustment (Figure 3A). When restricting the cohort to first born infants (N=7,541; 3,107 unexposed and 4,344 exposed), results from unadjusted, partially adjusted and adjusted models are consistent with those from the entire cohort (Figure 3B).

**Table 1.** Characteristics of mothers and infants in the maternal–paediatric cohort, by maternal COVID-19 vaccination status.

| Characteristic |  | Maternal COVID-19 vaccination status |  |  |
| --- | --- | --- | --- | --- |
|  |  | No protection<br>N = 4,868<sup>1</sup> | ≥1 dose before<br>delivery<br>N = 6,744<sup>1</sup> | Overall<br>N =11,612<sup>1</sup> |
| Maternal age at delivery, years |  | 33.32 (5.22) | 34.19 (4.90) | 33.82 (5.05) |
| Gestational age, weeks |  | 38.96 (1.87) | 38.91 (1.58) | 38.93 (1.72) |
| Birth country |  |  |  |  |
|  | Italy | 3,312 (68.0) | 5,263 (78.0) | 8,575 (73.8) |
|  | Other | 1,556 (32.0) | 1,481(22.0) | 3,037 (26.2) |
| Employment |  |  |  |  |
|  | Employed | 3,353 (68.9) | 5,384 (79.8) | 8,737 (75.2) |
|  | Unemployed | 505 (10.4) | 515 (7.60) | 1,020 (8.8) |
|  | Student | 30 (0.6) | 25 (0.4) | 55 (0.5) |
|  | Housewife | 976 (20.0) | 815 (12.1) | 1,791 (15.4) |
|  | Other | 4 (0.1) | 5 (0.1) | 9 (0.1) |
| Educational level |  |  |  |  |
|  | Low | 886 (18.2) | 815 (12.1) | 1,701 (14.6) |
|  | Intermediate | 2,095 (43.0) | 2,355 (34.9) | 4,450 (38.3) |
|  | High | 1,887 (38.8) | 3,574 (53.0) | 5,461 (47.0) |
| Singleton pregnancy |  |  |  |  |
|  | Yes | 4,697 (96.5) | 6,572 (97.4) | 11,269 (97.0) |
|  | No | 171 (3.5) | 172 (2.6) | 343 (3.0) |
| Firstborn infant |  |  |  |  |
|  | Yes | 3,197 (65.7) | 4,344 (64.4) | 7,541 (64.9) |
|  | No | 1,671 (34.3) | 2,400 (35.6) | 4,071 (35.1) |
| Premature birth |  |  |  |  |
|  | Yes | 396 (8.1) | 373 (5.5) | 769 (6.6) |
|  | No | 4,472 (91.9) | 6,371 (94.5) | 10,843 (93.4) |
| Sex |  |  |  |  |
|  | Male | 2,477 (50.9) | 3,522 (52.2) | 5,999 (51.7) |
|  | Female | 2,391 (49.1) | 3,222 (47.8) | 5,613 (48.3) |

<sup>1</sup>Mean (SD); n (%)

**Figure 3:**
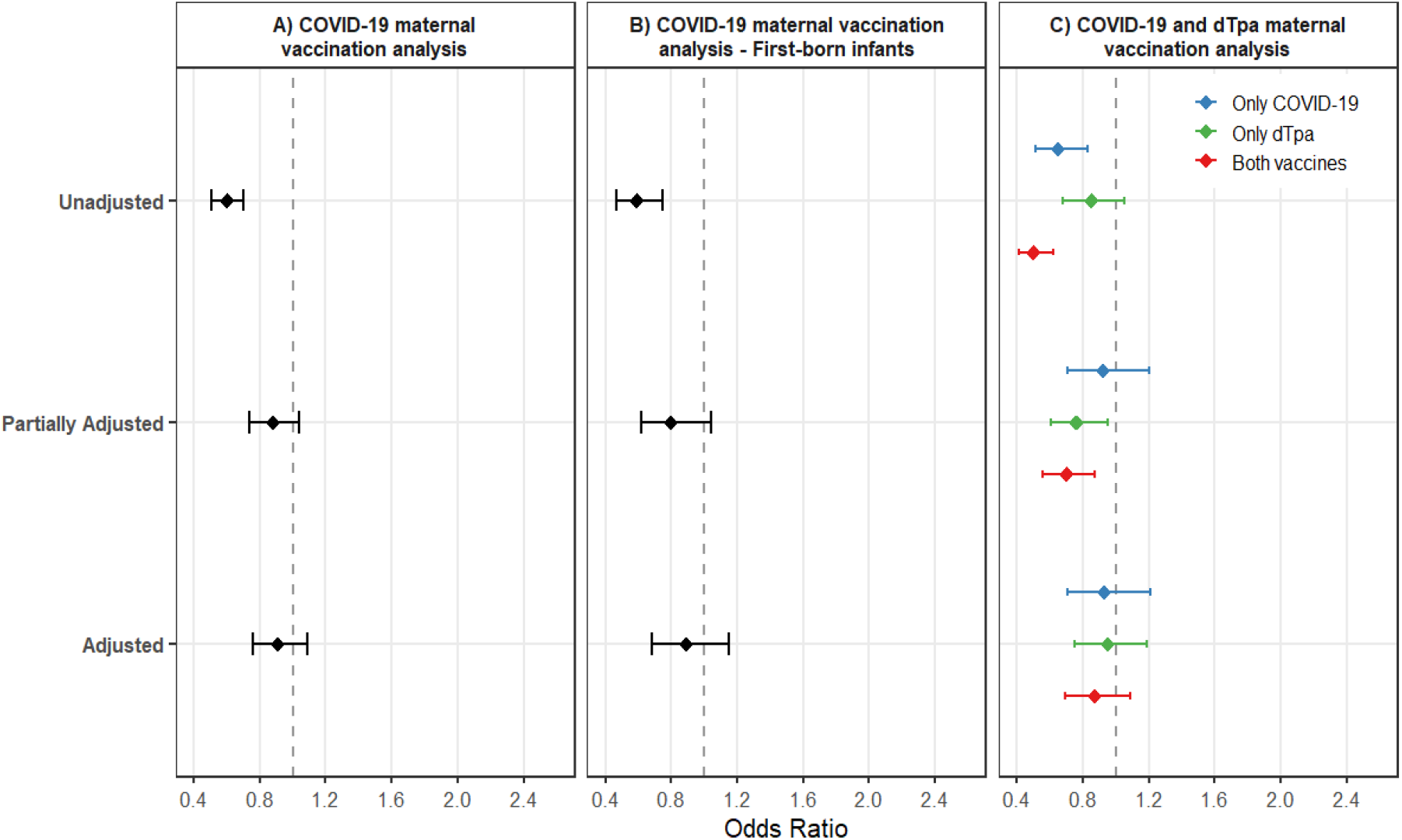
Forest plot of the association between maternal vaccination and infant bronchiolitis emergency room visits. Points represent odds ratios, and horizontal lines represent 95% confidence intervals. Estimates are shown for (A) maternal COVID-19 vaccination; (B) maternal COVID-19 vaccination in the analysis restricted to firstborn infants; and (C) maternal COVID-19 and DTaP vaccination. Results are presented for the unadjusted, partially adjusted, and fully adjusted models. The partially adjusted model includes categorical month–year of delivery and newborn sex. The fully adjusted model additionally includes maternal age at delivery, maternal country of birth (Italy or abroad), place of residence at delivery, employment status, educational level, and maternal history of gestational hypertension, chronic hypertension, pre-eclampsia, gestational diabetes, diabetes, deep-vein thrombosis or pulmonary embolism, antiplatelet use, oral anticoagulant use, lipid-lowering treatment, depression, and anxiety. The dashed vertical line indicates the null value (OR=1). DTaP=diphtheria, tetanus, and acellular pertussis. OR=odds ratio.

When evaluating specificity of SARS-CoV2 protection as compared to other maternal immunisations (Tdap), we found little support for a protective effect of either vaccinations on their own (COVID-19 vaccination only: OR=0.93 [0.71–1.20], Tdap vaccination only: OR=0.95 [0.75–1.19]), or jointly (OR=0.87 [0.69–1.09]) (Figure 3C).

### COVID-19 indirect effects

In terms of possible shifts in parental health seeking behaviour, hospitalised bronchiolitis consistently accounted for approximately half of the total incidence rate both before and after the pandemic (Figure 2). Multiple visits for the same disease episode played a minor role: when considering a 15-day washout period per episode, seasonal peaks were similarly attenuated across the entire period, with peaks in 2021–2022 and 2022–2023 approximately twice as high as pre-pandemic (Figure S2). Relative age distribution of bronchiolitis ER visits did not show substantial variation across seasons, indicating a stable pattern in the age at presentation (Figure S3).

A total of 103,494 infants was eligible for inclusion in the restricted paediatric cohort to explore impact of infant immunizations (delays in schedules, missing doses etc). Within this group, 97.3% of infants received at least one dose of the hexavalent or pneumococcal vaccine, with 77.9% vaccinated within the first 90 days of life. We found little evidence of association between early infant vaccination and ER visits for bronchiolitis (adjustedHR=1.09 [0.98–1.22]).

## Discussion

We describe a dramatic post-pandemic doubling in the burden of infant bronchiolitis in Northen Italy, rule out data artefacts as possible explanations, confirm a substantial increase in cumulative seasonal burden, and systematically investigate plausible drivers of this epidemiological shift, including the direct and indirect impacts of the COVID-19 pandemic. In the context of the recent introduction of highly effective RSV preventive strategies, infant immunization with Nirsevimab in Lombardy has already been associated with substantial reductions in RSV-related emergency department visits and hospitalizations^21^. However, understanding the factors driving the changing epidemiology of bronchiolitis is critical to maximise preventative public health impact and identify complementary prevention strategies. To address this, we combined population-wide individual-level healthcare data with deep phenotyping, including complete vaccination histories and detailed emergency department diagnoses. We found no or at best little evidence that changes in disease severity, repeated emergency department attendance, or infant and maternal vaccination patterns, including COVID-19 vaccinations, accounted for the observed increase. Taken together, our findings suggest that the post-pandemic rise in bronchiolitis reflects a complex interplay of factors affecting both hosts and healthcare systems, highlighting the need for continued epidemiological surveillance to optimize the implementation and evaluation of emerging RSV immunization programs.

Our temporal description—showing a substantially reduced incidence during the 2020-2021 season followed by a near-doubling of infant bronchiolitis cases in 2021-2023—aligns with global trends^6^. The near-zero incidence during the 2020–2021 season has been widely reported across multiple countries and is thought to be a consequence of stringent non-pharmaceutical interventions. Similar rebounds have been documented internationally^6^. Although a direct comparison of magnitudes is limited due to the use of different units of measurement across studies, the overall pattern of a heightened and temporally shifted RSV season is consistent. Our findings also align with the trends observed in several localized Italian studies^8–10^. These observations contrast with earlier, more localized reports from Rome that showed a slight decline in 2021-2022^7^, a discrepancy that may reflect differences in timing or study design. In addition, in our study the greater AUTS observed during the last two seasons confirms that the higher peak cannot be explained only by a change in the timing of seasonal activity. Instead, it reflects a genuine increase in the overall seasonal burden.

We find little evidence that maternal COVID-19 vaccination during pregnancy, used as an indirect proxy for prenatal and early-life SARS-CoV-2-related protection, was associated with infant bronchiolitis. SARS-CoV-2 infection has been hypothesised to increase susceptibility to subsequent respiratory infections by altering early-life immune development, following infant or maternal infection during pregnancy^15,22^. As direct evidence on early-life SARS-CoV-2 exposure was unavailable, we use maternal COVID-19 vaccination during pregnancy as a proxy for prenatal SARS-CoV-2-related immune protection, supported by evidence that maternal COVID-19 vaccination reduces SARS-CoV-2-related hospitalisations in early life^23^. We find no evidence that maternal COVID-19 vaccination is associated with bronchiolitis risk, and this conclusion in consistent across all sensitivity analysis. Restring the analysis to first-born infants does not change the results, suggesting that the presence of older siblings is unlikely to mask a protective effect of maternal vaccination. Similarly, when including Tdap vaccination, none of the maternal vaccination groups show a clear association.

Our results essentially rule out the hypothesis that recent inflated infant bronchiolitis rates reflect changes in healthcare provision (infant immunisation schedules or delivery) or parental health seeking behaviour. Although routine childhood immunisation was disrupted during the COVID-19 pandemic, with delays in vaccine delivery and modest reductions in vaccination coverage reported in Italy^24^, these factors were not associated with bronchiolitis risk in our analyses and therefore are unlikely to explain the observed trends. Regarding the hypothesis of a marked influence of increased parental caution, leading to a rise in emergency room visits for milder cases or increased in repeated presentations for the same case, we similarly find little support in our data, with stable proportions of ER admissions requiring hospitalisation and of episodes with multiple presentations throughout the analysis period.

An alternative hypothesis behind the post-pandemic rise related to the ‘immunity debt’, with some authors hypothesising that reduced exposure to microorganisms including RSV during pandemic lockdowns may have contributed to an increased number of cases compared to previous historical trends^25,26^. This mechanism would not directly affect the infant population in our study, who had not been born yet in 2020–2021, yet we cannot rule out an ‘immunity debt’ effect acting via reduced maternal RSV immunity and lower levels of antibodies transferred to their infants in the following epidemic season (2021-2022)^27^. However, such an effect would be expected to be strongest in the first post-pandemic epidemic season and is therefore unlikely to convincingly account for the sustained increase observed in 2022–2023, which remains unexplained.

This study has several strengths. First, we use population-level data from the entire Lombardy region, accounting for >15% of the national population. This represents a much larger sample size than previous region-or hospital-based studies. Because Lombardy is representative of the national context, our findings are generalisable to Italy and other European countries with similar healthcare systems. Second, the long observation period, (2012-2023) allows us to establish a robust pre-pandemic baseline and to follow the trends across two full RSV seasons after the COVID-19 pandemic. Third, the linkage of different administrative databases allowed us to test specific hypotheses that previous studies could not address. By connecting birth records (CeDAP) with ER visits and vaccination data, we included detailed information on both mothers and babies. This approach enabled us to evaluate the impact of various factors—such as maternal and infants vaccinations—on the risk of bronchiolitis. Finally, our systematic approach allowed us to distinguish between potential data artifacts and real epidemiological changes. By analysing metrics like ER and hospital admission rates or AUTS, we were able to provide a more reliable picture of the actual increase in the seasonal burden of RSV.

We acknowledge some limitations to the present analyses. First, bronchiolitis was used as a clinical proxy for RSV infection without laboratory confirmation, which may have led to some diagnostic misclassification. However, bronchiolitis in infants represents a robust clinical phenotype, strongly predictive of RSV infection (60-80% of cases are laboratory confirmed RSV cases in this age group^28^), and the proportion of RSV-positive tests among infants with bronchiolitis remained stable despite increased testing^29^. Thus, increased testing is unlikely to explain the observed increase in rates. Second, our outcome was restricted to ER visits; therefore, milder cases managed in primary care were not captured. This may underestimate the true community burden and potentially obscure the protective effects of vaccinations in less severe cases. Nevertheless, ER attendance represents clinically relevant bronchiolitis episodes consistently captured through population-wide registries, making it unlikely that exclusion of milder primary care-managed cases substantially biased the temporal trends. Moreover, currently available RSV preventive strategies have shown their substantial effects against medically attended and severe disease outcomes^30^, suggesting that ER visits are an appropriate outcome for evaluating potential protective effects. Third, because SARS-CoV-2 testing was not routinely performed in infants, we could not directly assess the contribution of COVID-19 infection in this age group. To address this limitation, we used maternal COVID-19 vaccination during pregnancy as a proxy for reduced early-life SARS-CoV-2 exposure and its consequences. Although this approach cannot fully capture individual infection history, the lack of association observed in adjusted analyses suggests that SARS-CoV-2 exposure alone is unlikely to explain the substantial increase in bronchiolitis ER visits. Fourth, we did not consider maternal influenza vaccination as putative protective factor due to extreme seasonality in vaccination availability and uptake resulting in skewed exposure distribution and substantial temporal confounding, making it difficult to disentangle potential vaccine effects from underlying seasonal viral circulation patterns. Fifth, despite our comprehensive dataset, some unmeasured confounders, such as breastfeeding duration, household crowding, and exposure to environmental tobacco smoke, may have influenced our assessment of the putative explanatory factors.

In conclusion, our study substantially contributes evidence to inform public health strategies to prevent infant RSV and bronchiolitis. We confirm a dramatic and sustained increase in infant bronchiolitis ER visits following the nadir of the 2020-2021 season in Lombardy. We rule out any single direct or indirect COVID-19 effect, including changes in disease severity, shape of the epidemic peak, parental health seeking behaviour and vaccine coverage. Immunity debt may have contributed to the first post-pandemic season but is unlikely to explain the persistence of elevated rates thereafter. Rather than a single factor, it is likely that we are witnessing a complex combination of multiple epidemiological shifts, such as altered viral dynamics, changes in social behaviours, and difficult-to-capture changes in healthcare provision and attendance. Identifying the mechanisms underlying these changes will be essential for developing effective and affordable public health strategies to reduce the burden of infant bronchiolitis and better anticipate future RSV epidemics.

## Contributors

VN, ANG, and LZ conceived and designed the study. LZ established the cohort and was responsible for data oversight and management. VN performed the data analyses and wrote the first draft of the manuscript. VN, ANG, LG, CG, AP, AB, and LZ contributed to the interpretation of the data and critically revised the manuscript for important intellectual content. All authors read and approved the final manuscript. VN and LZ were responsible for the decision to submit the manuscript.

## Data sharing

The data that support the findings of this study are available from Lombardy Region, but restrictions apply to the availability of these data, which were used under licence for the current study, and so are not publicly available.

Information on how to request access to Lombardy Region data is publicly available on the Epidemiological Observatory of Lombardy Region website (https://www.osservatorioepidemiologico.regione.lombardia.it/wps/portal/site/osservatorio-epidemiologico/DettaglioRedazionale/collaborazioni-con-gli-enti/daas+2-0/red-daas-2-0). Access may be requested by submitting a project application for individual-level data. The application should specify the purpose of data use; the requested data and variables; the definitions of the target and control groups; the requested study period; and the data use plan. Applications are evaluated on a case-by-case basis. Once approved, the data are made available within the secure DaaS 2.0 computing environment.

## Acknowledgments

This study was carried out according to the Lombardy Region laws on the use of regional healthcare databases for research activities (D.g.r. X/4893, 09/03/2016; D.g.r. XI/491, 02/08/2018; D.g.r. XI/6387, 16/05/2022.), and in particular on COVID-19 disease (D.g.r XI/3019, 30/03/2020). We thank Marco Villa and the Struttura Controllo e monitoraggio dati, LEA e outcome and the Azienda Regionale per l’Innovazione e gli Acquisti (ARIA) S.p.a. for the support of this research. We acknowledge support from the grant CEX2023-001290-S funded by MCIN/AEI/10.13039/501100011033, and support from the Generalitat de Catalunya, Spain, through the CERCA Program. ANG received funding from the European Union’s Horizon Europe research and innovation programme under the Marie Skłodowska-Curie Grant Agreement No. 101153708 (TinyTrend). The authors would also like to thank the data managers at Lombardy Region and Human Technopole for their helpful assistance. We are grateful to Francesca Ieva and Maeregu Woldeyes Arisido for their valuable feedback on earlier drafts of this manuscript.

