## Supplementary appendix for "Direct and indirect effects of COVID-19 on infant bronchiolitis: a population-wide registry study from Northern Italy"

**Contents**


1. **Supplementary tables**
   - Table S1: Description of the administrative healthcare datasets used in the study, including the key variables and their role in the analysis. [p. 2]
   - Table S2: Table S2: ICD-9 and ATC codes used to define confounders. [p. 3]
   - Table S3: AUTS for seasonal burden of ER visits for bronchiolitis. [p. 4]
2. **Supplementary figures**
   - Figure S1: Decreasing Person-Time Trend for Infants Aged 0–6 Months [p. 5]
   - Figure S2: Monthly incidence rates of bronchiolitis-related emergency room visits after applying a 15-day washout period following each ER visit. [p. 6]
   - Figure S3: Age distribution of infants presenting to the emergency department for bronchiolitis, 2012–2013 to 2022–2023 seasons. [p. 7]

**Table S1:** Description of the administrative healthcare datasets used in the study, including the key variables and their role in the analysis.

| **Dataset (original name)** | **English description** | **Variables used** | **Purpose in the study** |
|---|---|---|---|
| Anagrafica | Demographic information for women and infants in the study | cod_soggetto (identifier code);<br>anno_nascita (birth year or week-year);<br>nazione_nascita (country of birth);<br>stato_2012- stato_2023 (healthcare assistance status);<br>istat_dom_2012 – istat_dom _2023 (domicile); | Infants' identifier code, birth week year and healthcare assistance status were used to identify infants during the study period. Mother's identifier code, birth year, state of assistance and domicile were used in the maternal-paediatric cohort to complement information retrieved from CeDAP registry.<br>Healthcare assistance status was additionally used to verify continuous medical assistance in Lombardy according to the inclusion criteria. |
| Cedap_parti | Certificates of delivery assistance – mother's information | cod_soggetto (maternal identifier code);<br>cedap_key;<br>cond_prof_id_madre (maternal educational level);<br>data_parto (delivery date: year-week);<br>eta_gestazionale (gestational age);<br>titolo_id_madre (maternal education level) | The two datasets were linked using the CedAP key, enabling mother–child linkage for construction of the maternal–paediatric cohort. Because of missing values in children's identifier codes, linkage with the Anagrafica and other healthcare databases was not possible for all subjects. Delivery date was additionally used to validate information recorded in the Anagrafica dataset, while the remaining variables were included as covariates in the statistical models. |
| Cedap_figlio | Certificates of delivery assistance – mother's information | cod_soggetto (child's identifier code);<br>cedap_key;<br>data_parto (delivery date: year-week) | |
| PS | Emergency Room visits | cod_soggetto (identifier code)<br>id_accesso (visit identifier code)<br>diag1-diag2 (diagnosis)<br>dt_ingresso (admission date)<br>dt_dimissione (discharge date)<br>esito (visit outcome) | Identification of ER visits through ICD-9 diagnosis codes recorded in the diagnosis fields. The visit identifier was used to identify and remove duplicate records. Admission and discharge dates were used to determine the timing of each visit, while visit outcome was used to distinguish visits resulting in discharge from those leading to hospitalization. |
| Far_Terr | Drugs dispensations registry | cod_soggetto (identifier code)<br>atc (drug atc code)<br>dt_erogazione (dispensation date) | Identification of dispensed medications using Anatomical Therapeutic Chemical (ATC) codes and dispensation dates. These data were used to derive drug exposure variables included in the analyses. |
| Vaccinazioni | Vaccination registry | cod_soggetto (identifier code)<br>atc_id (vaccine atc code)<br>data_vaccino (vaccination date)<br>vaccino_id (vaccine acronym) | Identification of administered vaccines using ATC codes, vaccine acronyms, and vaccination dates. These data were used to derive infant and maternal vaccination status. |

**Table S2:** Table S2: ICD-9 and ATC codes used to define confounders. When searching for drugs, the first prescription is considered.

| Confounders | ICD-9 codes | ATC codes |
|---|---|---|
| History of gestational hypertension | 6423 | C02 C03 C07 C08 C09 |
| History of hypertension | 401 402 403 404 405 | C02 C03 C07 C08 C09 |
| History of preeclampsia | 6424 6425 6426 6427 6429 | // |
| History of gestational diabetes | 6488 | A10A A10B A10X |
| History of diabetes | 2500 2501 2502 2503 2504 2505 2506 2507 2508 | A10A A10B A10X |
| History of deep vein thrombosis or pulmonary embolism | 6713 6714 6719 451 452 453 673 4151 | // |
| History of antiplatelets | // | B01AC |
| History of oral anticoagulants | // | B01AE07 B01AF01 B01AF02 B01AA03 B01AA07 B01AF03 |
| History of lipid lower agents | // | C10A C10B |
| History of depression & anxiety | 3000 3001 3002 3003 3006 3007 3008 3009 3078 309 | N05BA N05CD N05BC01 N05BC51 N05BX N05CF N05CX01 N06BX |

**Table 3:** AUTS for seasonal burden of ER visits for bronchiolitis.

| ***Season*** | *2012-2013* | *2013-2014* | *2014-2015* | *2015-2016* | *2016-2017* | *2017-2018* | *2018-2019* | *2019-2020* | *2020-2021* | *2021-2022* | *2022-2023* |
|---|---|---|---|---|---|---|---|---|---|---|---|
| ***AUTS*** | *5.62* | *6.11* | *6.23* | *6.89* | *7.99* | *7.94* | *8.42* | *6.85* | *0.53* | *8.87* | *12.22* |

AUTS values quantify the seasonal burden of emergency room visits for bronchiolitis among infants aged 0–6 months across the eleven seasons included in the study (2012–2013 through 2022–2023). AUTS=area under the time series. ER=emergency room.

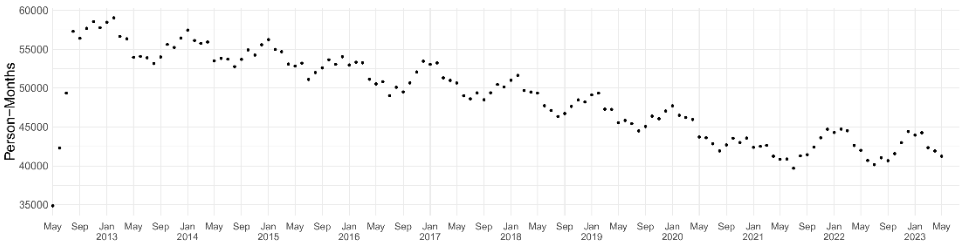


**Figure S1**: Decreasing Person-Time Trend for Infants Aged 0–6 Months. Trend in cumulative person-time for the 908,324 infants (age 0-6 months) included in the paediatric cohort in Lombardy. The data show a consistent decline in person-time over time, reflecting a reduction in the eligible study population size.

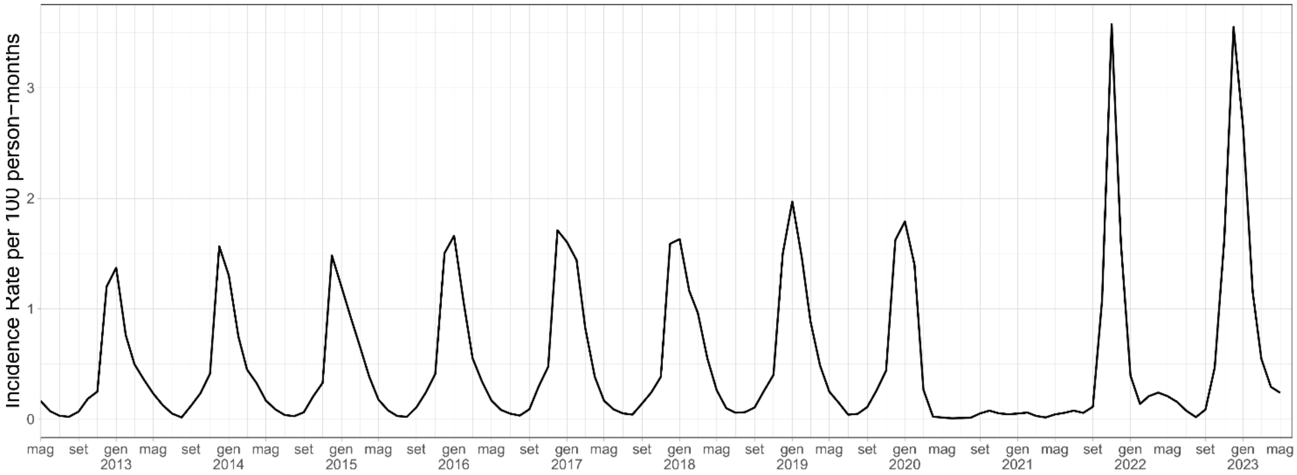


**Figure S2**: Monthly incidence rates of bronchiolitis-related emergency room visits after applying a 15-day washout period following each ER visit.

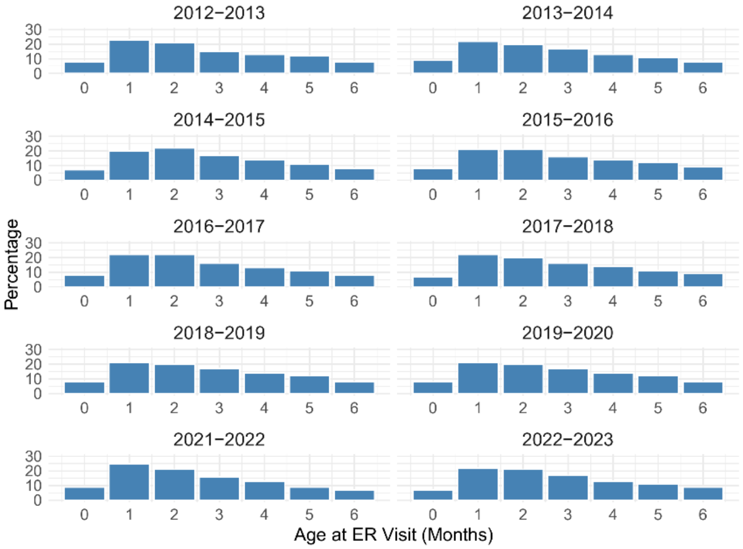


**Figure S3:** Age distribution of infants presenting to the emergency department for bronchiolitis, 2012–2013 to 2022–2023 seasons (2019–2020 season excluded due to low number of cases)
